# Self-reported Morisky 8 item medication adherence scale to statins concords with pill count method and correlates with serum lipid profile parameters and Serum HMGCoA Reductase levels

**DOI:** 10.1101/19006148

**Authors:** Abhinav Grover, Mansi Oberoi, Harmeet Rehan, Lalit Gupta, Madhur Yadav

## Abstract

**Background:** It is imperative that non-compliance to statins be identified and addressed to optimize the clinical benefit of statins. Patient self-reporting methods are convenient to apply in clinical practice but need to be validated.

**Objective:** We studied the concordance of a patient self-report method, MMAS (Morisky eight item medication adherence scale) with pill count method in measuring adherence to statins and their correlation with extended lipid profile parameters and serum HMGCoA-R (hydroxymethylglutaryl coenzyme A reductase) enzyme levels.

**Methods:** MMAS and pill count method were used to measure the adherence to statins in patients on statins for any duration. Patients were subjected to estimation of extended lipid profile and serum HMGCoA-R levels at the end of 3 months follow-up.

**Results:** Out of a total of 200 patients included in the study, 117 patients had low adherence (score less than 6 on MMAS) whereas 65 and 18 patients had medium (score 6 to less than 8) and high adherence (score of 8) respectively. Majority of patients who had low adherence to statins by MMAS were nonadherent by pill count method yielding concordance of 96.5%. Medium or high adherence to statins by MMAS method had concordance of 89.1% with pill count method. The levels of total cholesterol, low density lipoprotein-cholesterol, apolipoprotein B and HMGCoA-R were significantly negatively correlated with compliance measured by pill count and MMAS with similar correlation coefficients. HMGCoA-R levels demonstrated a plateau phenomenon with levels being 9-10 ng/ml when compliance to statin therapy was greater than 60% by pill count and greater than 6 on Morisky scale.

**Conclusion:** In conclusion, MMAS and pill count methods showed concordance in measuring adherence to statins. These methods need to be explored further for their interchangeability as surrogates for biomarker levels.

## INTRODUCTION

Statins are the most effective lipid-lowering agents which reduce the risk of coronary events by 17%-26%. (1) The benefits of statins are lost when patients are poorly compliant and it is reported that only 50% of patients who are being treated with statins continue to use their medication after six months, and only 30% to 40% after one year. (2, 3) Hence, it is imperative that non-compliance to statins be identified to optimize the clinical benefit of statins. Usually, treating physicians do not inquire about patients’ adherence to the medication (4). Medication adherence can be assessed by pill count, pharmacy fill rates, surrogate marker levels, patient self-reporting methods e.g. Morisky medication adherence scale 8-item (MMAS) etc. The most convenient ones are patient self-reporting methods which can be easily applied in clinical setting but they are generally not specific for a disease or drug. (6, 7) There is a need to understand and validate MMAS for measuring compliance to statins in dyslipidemic patients. Hence, the present study evaluated the concordance of MMAS(8) with pill count method in measuring compliance to statins in dyslipidemic patients and also correlated the extent of compliance to statins with lipid profile like and serum 3-hydroxyl-3-methylglutaryl coenzyme A reductase (HMGCoA-R) enzyme levels.

## MATERIALS AND METHODS

### Subjects

Dyslipidemic patients with age above 18 years, elevated LDL (low density lipoprotein) levels and/or TG (triglyceride) levels, and/or low HDL (high density lipoprotein) levels as per ACC-AHA (American College of Cardiology/ American Heart Association) guidelines (2) and on statin therapy for any duration were included in the study. Patients who had ACS (acute coronary syndrome) within the last 3 months, history of hypothyroidism, pregnancy/lactation and hypersensitivity or intolerance to statins were excluded from the study.

### Study Design

In a prospective observational study, information of patients’ personal, demographic and socioeconomic status was recorded. They were assessed for compliance to statins using MMAS and pill count method (9) at the end of 3 month after refilling of medicine. Patients with a score of pill count ≥80% were considered compliant whereas based on MMAS scores, the patients were categorized into the 3 levels of adherence i.e. high adherence (score, 8), medium adherence (score, 6 to <8), and low adherence (score, <6) to facilitate its use. (8) (Figure 1)

**Figure 1:**
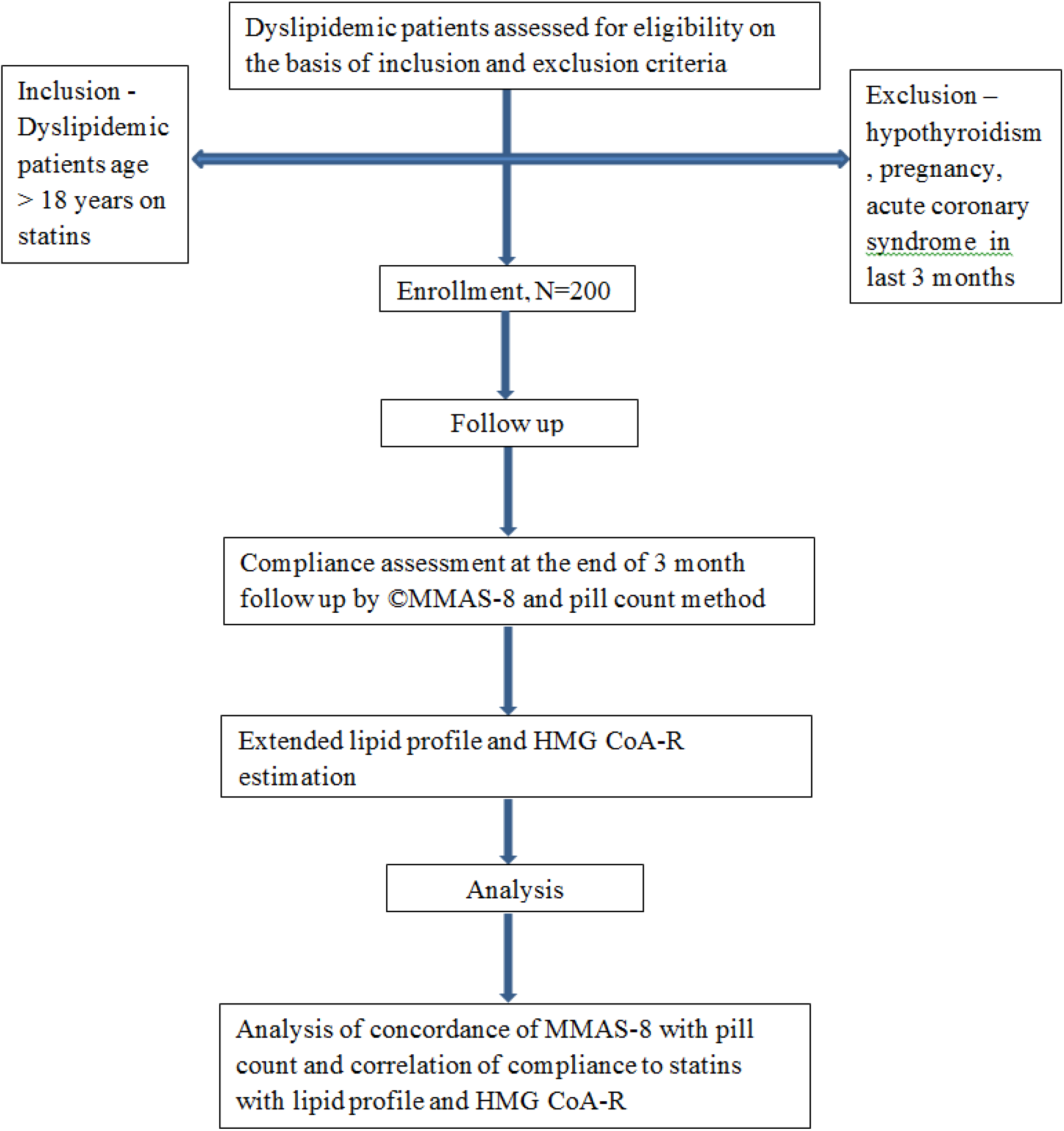
Study design. HMG CoA-R = 3-hydroxyl-3-methylglutaryl coenzyme A reductase ; MMAS = Morisky Medication Adherence scale

Study protocol was approved by the Institutional Human Ethical Committee. A written informed consent was taken from all the patients.

The decision to start the statins or to escalate their doses if required was at the discretion of the treating physician. Patients who required dose modification from one intensity of statin therapy to another with in the follow up period were excluded from the study.

A 5 ml venous blood sample of all the patients was taken to analyse extended lipid profile parameters including total cholesterol (TC), LDL, TG, HDL, Apolipoprotein A1, Apolipoprotein B and serum HMG CoA-R enzyme levels using ELISA (enzyme linked immunosorbesnt assay) method with BMAssay HMGCoA-R kit and AssayPro Apo A1 and Apo B ELISA kits.

### Statistical Considerations

The compliance data are presented as percentages for pill count and as mean of proportions for MMAS whereas lipid profile parameters are presented as mean±standard deviation and the agreement between MMAS and pill count is described by percentage concordance. The trend of pill count across MMAS categories was analysed using least square and maximum likelihood ratio for discrete and continuous variables respectively. The Pearson’s correlation analysis was used for correlation for compliance with lipid profile and serum HMG CoA reductase levels. P <0.05 was considered statistically significant.

## RESULTS

Out of a total of 200 patients included in the study 101 (50.5%) were females. The overall mean age of all the patients was 55.15±10.23 years (range, 23 to 82 years). The mean duration of prescription for statin at the time of enrollment was 8.6±13.08 months (range, 1-72 months). The demographic and clinical characteristics of patients by low, medium and high adherence MMAS category are described in Table 1.

**Table 1:** Table 1-Patients’ demographic and clinical characteristics as per MMAS category

|  | MMAS category ( score range) |  |  | P value |
| --- | --- | --- | --- | --- |
|  | Low (<6)<br>(N=117) | Medium (6 to<br><8) (N=65) | High (8) (N=18) |  |
| Age (years)<br>(Mean ± S.D) | $54.7 \pm 9.9$ | $55.9 \pm 11.1$ | $55.3 \pm 11.5$ | 0.82 |
| Gender<br>Females, N (%) | 59 (50.4) | 32 (49.2) | 10 (55.5) | 0.86 |
| **Comorbid conditions** |  |  |  |  |
| Diabetes mellitus,<br>N (%) | 76 (64.9) | 46 (70.7) | 14 (77.7) | 0.50 |
| Hypertension,<br>N (%) | 60 (51.2) | 31 (47.7) | 4 (22.2) | 0.07 |
| Ischemic<br>Heart disease, N<br>(%) | 11 (9.4) | 4 (6.2) | 1 (5.5) | 0.71 |

Low adherence to statins by MMAS method was observed in 117 patients who also showed no adherence by pill count method (n=113) yielding concordance of 96.5%. There were 83 patients who had medium or high adherence to statins by MMAS method and 74 of these were adherent by pill count method yielding concordance of 89.1%. When low and medium adherence categories by MMAS were clubbed together, their concordance with nonadherence by pill count was 67% whereas the concordance of high adherence by MMAS with adherence by pill count was 100%. (Table 2)

**Table 2:** Comparison of statin compliance by MMAS and pill count method

| Parameters | Pill count | MMAS category ( score range) |  |  | p value |
| --- | --- | --- | --- | --- | --- |
|  | method | Low (<6) | Medium (6 to <8) | High (8) |  |
| Number of patients (N) | 200 | 117 | 65 | 18 |  |
| Mean pill count (mean±SD) | 56.71 | 45.10±10.69 | 68.42±9.46 | 89.87±3.16 | **0.000** |
| Number of non-adherers (pill count <80%)(N) | 121 | 113 (96.58%) | 9 (13.84%) | 0 (0%) | **0.000** |
| Number of adherers (pill count ≥80%)(N) | 79 | 4 (3.41%) | 56 (86.15%) | 18 (100%) | **0.000** |

The levels of total cholesterol, LDL, HMG CoA-R and Apo B were significantly negatively correlated with compliance measured by pill count and MMAS. The levels of HDL-C were positively correlated with compliance by both measures. Serum LDL levels show similar negative correlation with compliance by MMAS (r=-0.750, p=0.000) and pill count (r=-0.776, p=0.000). HMG CoA-R also shows similar negative correlation with compliance by MMAS (r=- 0.497, p=0.000) and pill count (r=-0511, p=0.000). And Apo B shows negative correlation with MMAS (r=-0.239, p=0.001) and pill count (-0.233, p=0.001). (Table 3)

**Table 3:** Correlation of compliance by MMAS-8 and pill count method with extended lipid profile and HMGCoA-R.

| Method to study adherence |  | TC | TG | LDL | HDL | HMGC oA-R | ApoA | ApoB |
| --- | --- | --- | --- | --- | --- | --- | --- | --- |
|  |  |  |  |  |  |  | 1 |  |
| **MMAS-8** | Pearson Correlation | -0.643 | -0.487 | -0.750 | 0.781 | -0.497 | -0.025 | -0.239 |
|  | Sig. (2-tailed) | **0.000** | **0.000** | **0.000** | **0.000** | **0.000** | 0.727 | **0.001** |
| **Pill count** | Pearson Correlation | -0.602 | -0.498 | -0.776 | 0.785 | -0.511 | -0.039 | -0.233 |
|  | Sig. (2-tailed) | **0.000** | **0.000** | **0.000** | **0.000** | **0.000** | 0.583 | **0.001** |

HMG CoA-R levels demonstrated a plateau phenomenon with levels being 9-10 ng/ml when compliance to statin therapy was greater than 60% by pill count and greater than 6 on the Morisky scale whereas, the LDL-C levels achieved were 60-80 mg/dl with increase in compliance beyond these levels. (Figure 2-5)

**Figure 2:**
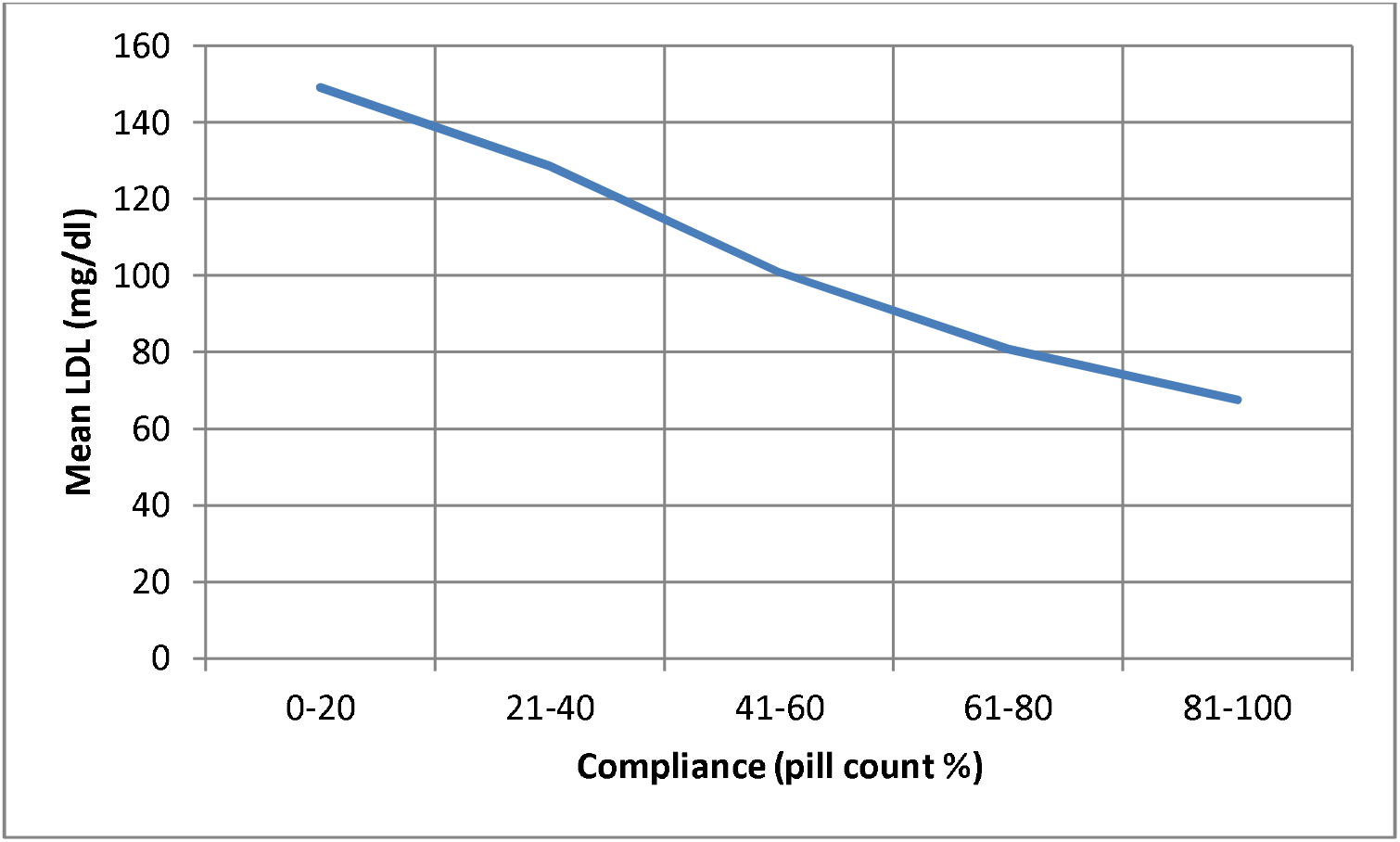
Plot of mean serum LDL levels against compliance measured using pill count.

**Figure 3:**
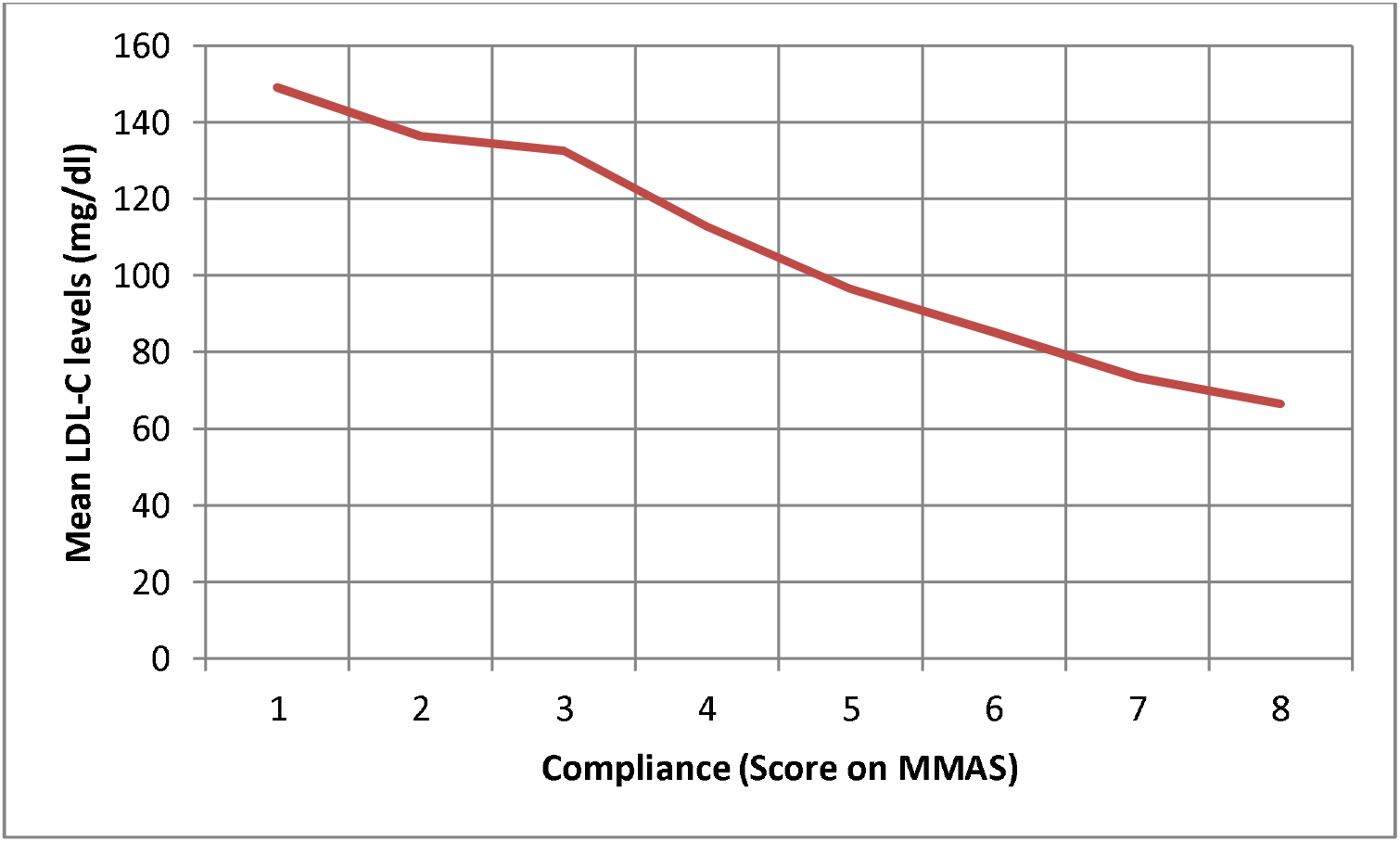
Plot of mean serum LDL levels against compliance measured using MMAS.

**Figure 4:**
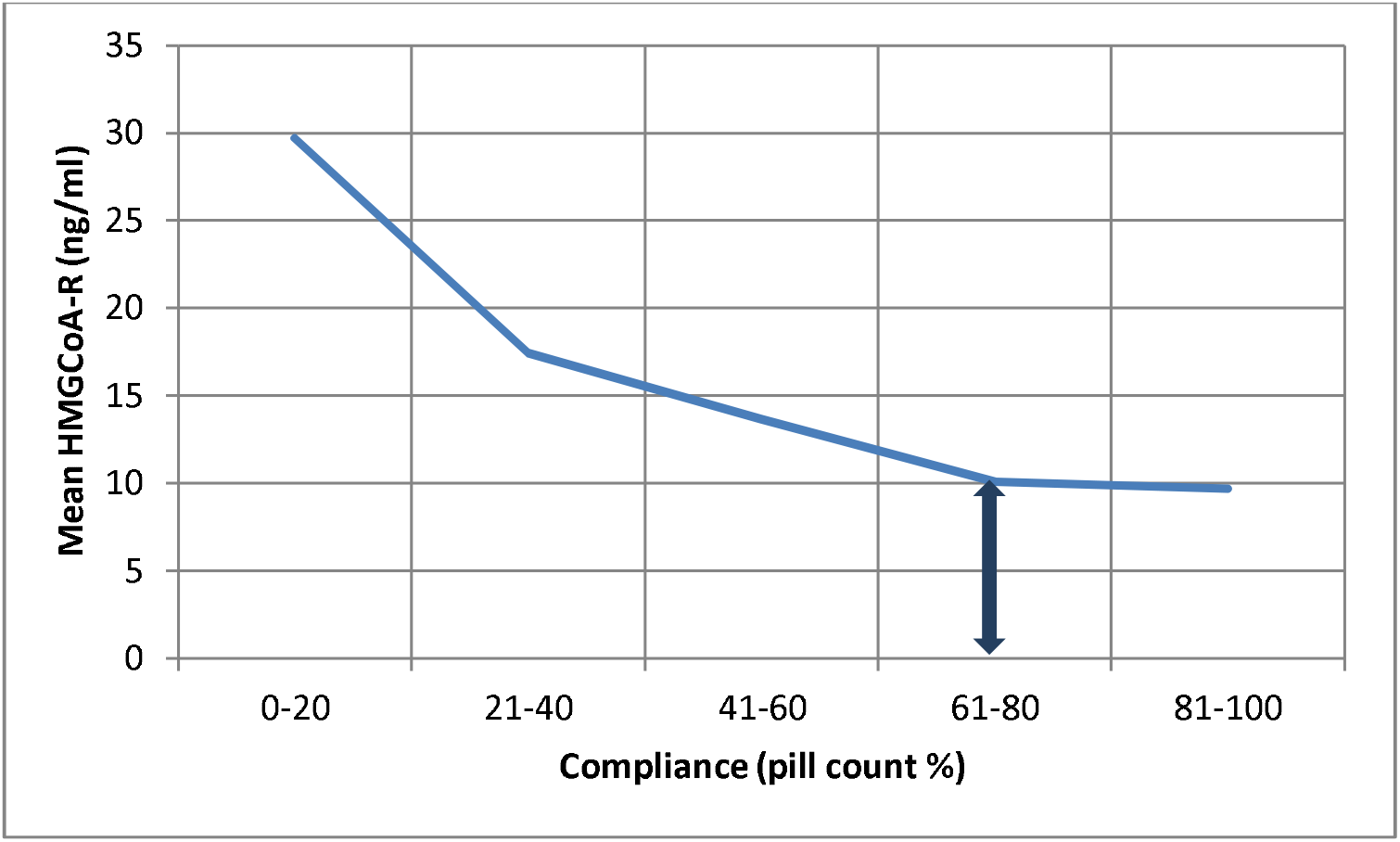
Plot of mean serum HMG CoA-R levels against compliance measured using pill count.

**Figure 5:**
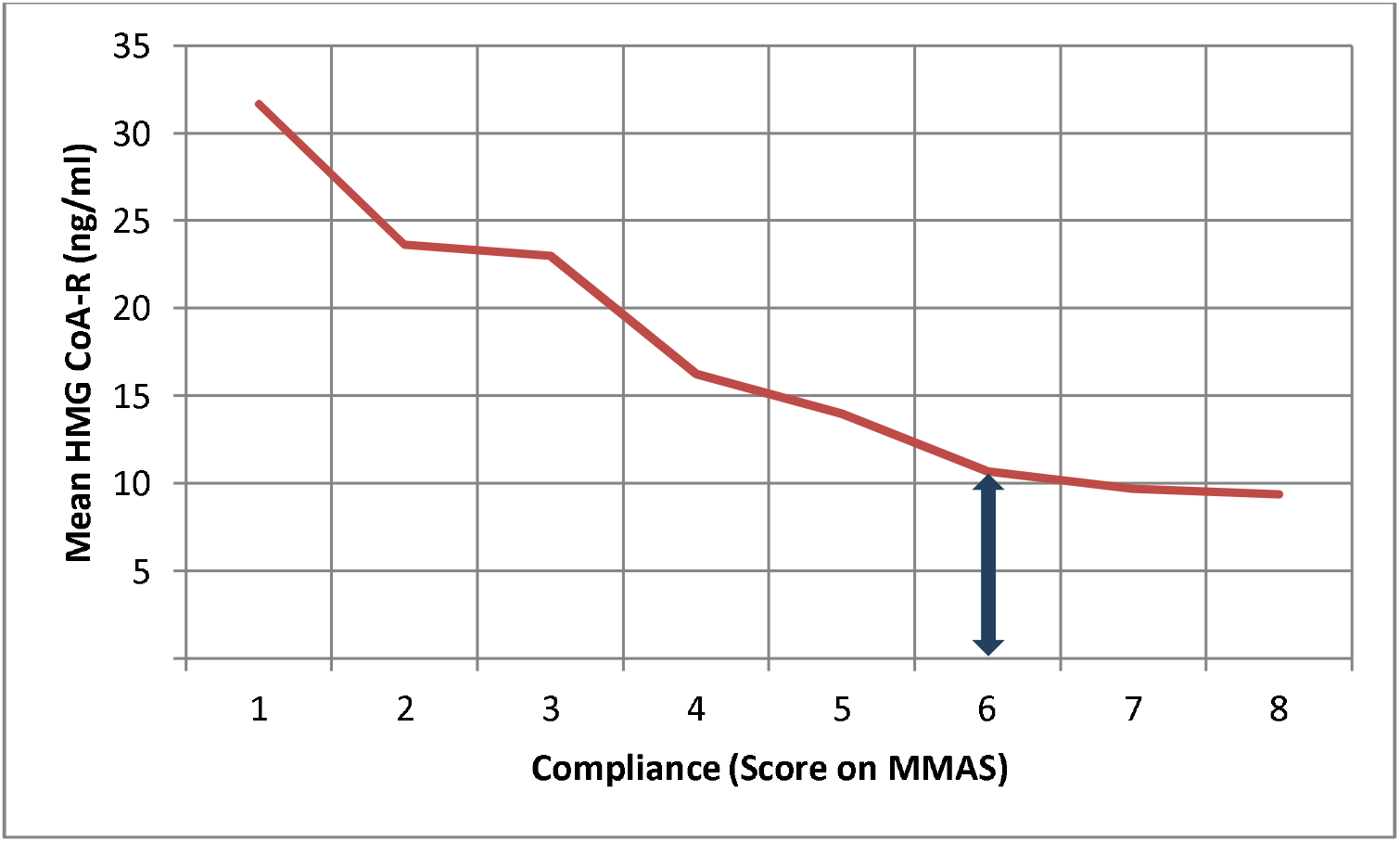
Plot of mean serum HMG CoA-R levels against compliance measured using MMAS.

## DISCUSSION

### Prevalence of non-compliance to statins

Compliance to statins is only around 50% at the end of 1 year of initiation of treatment. (10) Clinicians require information on medication adherence to draw proper conclusions about the effectiveness of treatment. The goal is to have access to a quick, reasonably accurate self-report adherence measure for use in outpatient settings to facilitate clinical decision making. A quick and accurate assessment of compliance can be especially beneficial for dyslipidemic patients in choosing between measures to increase compliance or switching to higher intensity of statins. (11)

### Compliance assessment for statins using Morisky medication adherence scale and pill count methods

We evaluated the association and concordance of the MMAS with pill count in measuring compliance to statins as no other study is available which evaluated this aspect. Majority of patients who had low adherence to statins by MMAS method were nonadherent by Pill count method showing concordance whereas medium or high adherence to statins by MMAS method yielded concordance of 89.1%. (Table 2). When low and medium adherence categories by MMAS were clubbed together, their concordance with nonadherence by pill count was 67% whereas the concordance of high adherence by MMAS with adherence by pill count was 100%. (Table 2) This suggested a score of lower than 6 on MMAS is a better predictor of non-adherence than defining non adherence as having a score of less than 8 on MMAS.

The identification of patients with low adherence may facilitate implementation of suitable interventions and aid in optimization of therapeutic benefit. There are numerous behaviors related to non-adherence which may be modifiable including lack of mindfulness, forgetting, complexity of treatment regimen etc. (12-20) Some of these can be identified using the responses of patients to individual items of MMAS and can enable appropriate interventions to improve adherence but this aspect needs to be explored further in dyslipidemic patients. (21)

### Compliance to statins and correlation with biomarkers of dyslipidemia

We also correlated the compliance to statins derived by MMAS and pill count methods with lipid profile and serum HMG CoA reductase enzyme levels. There was a significant negative correlation between total cholesterol, LDL, HMG CoA-R and Apo B levels and compliance by pill count and MMAS. Also, HDL shows significant positive correlation with compliance by both methods. Serum LDL levels show similar negative correlation with compliance by MMAS (r=-0.750, p=0.000) and pill count (r=-0.776, p=0.000). And Apo B also shows similar negative correlation with MMAS (r=-0.239, p=0.001) and pill count (-0.233, p=0.001) No study is available to compare the findings of our study. The similar correlation coefficients of different parameters with compliance by MMAS and Pill count suggest a parallel in MMAS and pill count methods as measures of compliance.

HMG CoA-R shows similar negative correlation with compliance by MMAS (r=-0.497, p=0.000) and pill count (r=-0511, p=0.000). Also, the levels of HMG CoA-R show a plateau at levels of 9-10 ng/ml when compliance is beyond 60% by pill count method and greater than score of 6 on MMAS. (Figure 4, 5) This suggested that high adherence defined by either of the two methods correlates with HMG CoA-R level of 9-10 ng/ml and this cut off could potentially be explored as a surrogate marker for defining adherence and non-adherence. Also, it would be beneficial in estimating the effectiveness of treatment.

### Study Limitations

Shortcomings of self-report include reliance on recall (Questions 1,2,4 and 5 in MMAS) and social desirability bias, with a tendency to overestimate adherence.(12) As an indirect measure, pill counts can misrepresent adherence as well, particularly since they fail to measure whether medication was taken on schedule.(9)

## CONCLUSIONS

MMAS and pill count methods of estimating compliance showed concordance and compliance by both the methods correlated in a similar manner with the modification in the extended lipid profile levels and with the inhibition of HMG CoA-R levels. Settings where lipid profile and HMG COA-R estimation is not available, if compliance of statins by MMAS is more than score of 6 or more than 60-80% by pill count method, it can be assumed that the benefit of statins have been adequately extended to the patient.

### Perspectives

## COMPETENCY IN MEDICAL KNOWLEDGE

Settings where lipid profile and HMG CoA-R estimation is not available, if compliance to statins by MMAS is more than score of 6 or more than 60-80% by pill count method, it can be assumed that the benefit of statins has been adequately extended to the patient.

## TRANSLATIONAL OUTLOOK

Future studies are needed to explore the utility of responses to individual items on Morisky medication adherence questionnaire to identify modifiable behaviors related to non-adherence to statins which can enable appropriate interventions to optimize therapeutic benefit.

## Data Availability

NA

## Acknowledgements

We thank Dr Morisky for permission of use of ©MMAS. Use of the ©MMAS is protected by US copyright laws. Permission for use is required. A license agreement is available from: Donald E. Morisky, ScD, ScM, MSPH, Professor, Department of Community Health Sciences, UCLA School of Public Health, 650 Charles E. Young Drive South, Los Angeles, CA 90095-1772.

